# Integrating Genome-wide and Epigenome-wide Associations for Antipsychotic Induced Extrapyramidal Side Effects

**DOI:** 10.1101/2025.02.27.25323006

**Authors:** Kai Yao, Johan H. Thygesen, Siobhan K. Lock, Antonio F. Pardiñas, Antonia L. Pritchard, Michael C. O’Donovan, Michael J. Owen, James T. R. Walters, David St Clair, Nick Bass, Andrew McQuillin

## Abstract

**Background and Hypothesis:** Antipsychotic medications are the first-line treatment for schizophrenia. However, around 40% of people with schizophrenia who are treated with antipsychotics could develop extrapyramidal side-effects (EPSE) including: 1) Dyskinesias, 2) Parkinsonism, 3) Akathisia, and 4) Dystonia.

**Study Design:** We conducted Genome-wide association (GWAS) and Epigenome-wide association (EWAS) meta-analysis of EPSE utilising data from previous schizophrenia case control studies. We integrated significant EWAS findings to an EPSE GWAS meta-analysis to enhance our understanding of the functional impact of common variants on EPSE. We also investigated whether polygenic risk scores (PRS) for schizophrenia, Parkinson’s disease, and Lewy-body dementia could be predictive of EPSE development.

**Study Results:** The top index SNP rs2709733 (A/G) from EPSE GWAS (p=2.214×10^-7^) mapped to a long intergenic non-protein coding RNA, *LINC01162* with consistent effects across all cohorts. We identified 9 differentially methylated positions (DMPs) associated with EPSE when controlling for methylation age, sex, derived estimates of cell composition, smoking score, and schizophrenia PRS. Four of the DMPs cg14531564, cg20647656, cg12004641, cg22845912, and their affiliated genes (*SDF4, ANKMY1, TNS1, SLA*) were associated with the risk of developing EPSE and not with schizophrenia risk. Another DMP (cg12044923) which mapped to the *STK32B* gene, showed significant enrichment for association with risk of EPSE.

**Conclusions:** Our study sheds new light on the potential biological mechanisms underlying EPSE development in schizophrenia, highlighting the importance of exploring both methylation shifts and common SNP associations. Further research with larger samples sizes and a focus on the role of *STK32B* are encouraged.

## Introduction

Antipsychotic medications are the first-line treatment for schizophrenia.^1^ Although many people benefit, around 70% may experience treatment failure such as psychiatric rehospitalization, suicide attempt, discontinuation or switch to other medication.^2^ Extrapyramidal side-effects (EPSE) are common with antipsychotic treatment,^3,4^ with approximately 40% of patients treated with first-generation antipsychotics (FGAs) experiencing EPSE.^5^ FGAs are primarily dopamine D2 receptor antagonists, which reduce dopaminergic activity to alleviate positive symptoms of psychosis. This, however, often leads to motor side effects such as EPSE,^6^ which describe the movement abnormalities induced by antipsychotics including:

1. **Dyskinesia**, hyperkinetic choreiform involuntary movements of the face, extremities, and the trunk.^7^ When dyskinesia persists for more than one month it is termed tardive dyskinesia which can sometimes become chronic.
2. **Parkinsonism**, symptoms of rigidity, tremor and impaired or slow movement (bradykinesia).^8^
3. **Akathisia**, characterised by subjective inner restlessness and objective increase in motor activity such as pacing.^9^
4. **Dystonia**, characterised by sustained and abnormal contractions, that can result in abnormal movements and postures.^10^

These movement abnormalities can lead to severe impairment and reduction in the quality of life of individuals with schizophrenia,^11^ by interfering with daily living activities and social functioning.^12,13^ In a meta-analysis, the prevalence of spontaneous dyskinesias and parkinsonism was found to be higher in antipsychotic-naive patients with schizophrenia and in first-degree relatives of patients with SCZ as compared to healthy controls, indicating a heritable, non-drug induced component to these abnormalities.^14^

Parkinsonism seen in EPSE can be clinically indistinguishable from the movement abnormalities seen in the neurological disorders like Parkinson’s disease (PD) and Lewy Body Dementia (LBD). Previous studies have identified shared significant loci between SCZ and PD.^15,16^ For example, SCZ and PD are both associated with the 22q11.2 deletion syndrome.^17^ A duplication of the *SNCA* gene, for which pathogenic variants are associated with autosomal dominant Parkinson’s and encodes α-synuclein, a major constituent of LBD, was reported in an individual diagnosed with SCZ nine years prior to the development of mild Parkinsonism.^18^ A recent neuroimaging study on individuals with first episode psychosis found that higher iron loading in the basal ganglia correlated with greater motor abnormalities including EPSE.^19^ Similar associations were found with motor abnormalities in PD.^20,21^ In view of this, it is plausible that there are shared genetic features between these disorders which also contribute to the shared phenotypical features including movement abnormalities like EPSE in SCZ.

Genome-wide Association Studies (GWAS) are a promising approach to identify potential genes associated with development of EPSE given the often-complex biological pathways implicated in psychiatric traits.^22^ However, to our knowledge, only one past study investigated antipsychotic induced EPSE using GWAS.^23^ The genotype data in that study had somewhat limited genomic coverage compared to contemporary studies and furthermore there was no imputation of genotypes not captured on the genotyping array. Epigenome-wide Association Study (EWAS) allows for the examination of environmentally induced methylome variation which could directly result from chronic antipsychotic exposure.^24,25^ To date, there has been no EWAS on EPSE to examine the influence from antipsychotics.

Our understanding of the molecular mechanisms underlying EPSE may be improved using an integrated functional genomics strategy. The overall aim of this study was to conduct an integrated GWAS and EWAS meta-analysis of EPSE data from existing schizophrenia studies. We also investigated whether Polygenic Risk Scores (PRS) for schizophrenia, PD and LBD could be used to predict risk of the development of EPSEs. The findings could provide a better understanding of the genetic underpinnings of EPSE and pave the way for the identification of informative genetic biomarkers that could allow for specific tailoring of treatments in the future.

## Method

### Participants Selection and Genotyping

#### UCL Participants

All UCL participants received an ICD10 diagnosis of schizophrenia from a UK National Health Service (NHS) psychiatrist. Details have been reported elsewhere.^26,27^ Ancestrally matched healthy controls were recruited from the National Health Service (NHS) blood transfusion service and from study sites where case participants were also being recruited. The healthy controls were screened for an absence of a lifetime history of the following disorders: schizophrenia and any other psychosis, major affective or schizoaffective disorders, eating disorders, alcohol/drug addiction, and obsessive-compulsive disorders. All participants read an approved information sheet and signed a physical informed consent form. The study was approved by the NHS Metropolitan Multi-centre Research Ethics Committee (MREC/03/11/090). Genome-wide single nucleotide polymorphism data were generated in three waves at the Broad Institute, Boston, MA, US, using the, Affymetrix Array, Illumina PsychArray, and Illumina Global Screening Array (GSA). The three waves of data underwent equivalent quality control and imputation methods which had been described in details elsewhere.^28^

### Aberdeen Participants

The Aberdeen case–control sample has been described elsewhere.^29^ Briefly, the cohort contains participants with schizophrenia and healthy controls who have self-identified as born in the British Isles (95% in Scotland). All participants with schizophrenia met the Diagnostic and Statistical Manual for Mental Disorders fourth edition (DSM-IV)^30^ and ICD-10 criteria for schizophrenia.^26^ Controls were volunteers recruited through general practices in Scotland. Volunteers who replied to a written invitation were interviewed using a short questionnaire to exclude major mental illness in the individual themselves and their first-degree relatives. The study was approved by both local and multiregional academic ethical committees and all cases and controls gave informed consent. The samples were genotyped at the Broad Institute, as described for the UCL participants.

### Cardiff Participants

Participants were recruited from community mental health teams in Wales and England on the basis of a clinical diagnosis of schizophrenia or schizoaffective disorder (depressed sub-type) as described previously.^31^ Diagnosis was confirmed following a SCAN interview^32^ and review of case notes followed by consensus diagnosis according to DSM-IV criteria^30^. The UK Multicentre Research Ethics Committee (MREC) approved the study and all participants provided informed consent. The samples were genotyped at the Broad Institute, as described for the UCL participants.

### UK Biobank (UKB) Participants

UKB is a biomedical database and research resource of approximately 500,000 individuals from across the UK aged 40 to 69 years at recruitment (between 2006 and 2010).^33^ Potential participants in UKB were selected using diagnosis of schizophrenia from ICD10, including codes from F20.0 to F20.9 and excluding participants with any primary Parkinson disorder with G20.

### Coding of Extrapyramidal Side-effects Data

Participant EPSE status was derived from clinical data recorded following: (1) diagnosis of schizophrenia, (2) prescription of antipsychotic medications (FGA or second generation antipsychotics; SGA); (3) recorded clinical features of EPSE side-effects; and/or recorded medications prescribed to alleviate EPSE side-effects. We used keywords to classify participants with schizophrenia as cases (having EPSE) or controls (not having EPSE). These key words covered two main areas: behavioural and pharmacological (Supplementary Table 1 and 2):

*1) Behavioural features of the four types of EPSE, (dystonia, akathisia, parkinsonism, and dyskinesia)*. To compile a list of keywords for each of these EPSE types, we consulted several rating scales that are frequently employed to measure EPSE including: The Abnormal Involuntary Movement Scale (AIMS),^34^ the Extrapyramidal Symptom Rating Scale (ESRS),^35^ The Simpson Angus Scale,^36^ and the Barnes Akathisia Rating Scale (BARS).^37^ In addition, we searched reliable sources of clinical information for each of these abnormalities including the National Institute for Health and Care Excellence (NICE) guidelines^38^ and the BMJ Best Practice.^39^
*2) Pharmacological treatments for EPSE.* To generate key words for pharmacological treatments for EPSE, we searched The NICE guidelines^38^ and The Maudsley Prescribing Guidelines in Psychiatry^40^ for the most recent recommendations on managing EPSE to identify a list of medications.

The UCL and Aberdeen participants’ EPSE status was derived using the same list of key words described in Supplementary Table S1 and S2. The Cardiff participants’ EPSE status coding had a few minor adaptions. The keywords “dribbling” was added as it better captured other saliva-related key-words; ‘shakes’ was removed as it was described in the context of anxiety; “still” was removed as it referred to still doing something not being physically still; “tap” was removed as it was in the context of ‘tapered’; “march” was removed as it referenced the month of March; “irritable” was removed as it was in the context of IBS/irritable bowel syndrome; “parkin” was removed as it referred to Parkinson’s disease not parkinsonism; ‘tropin’ was excluded as it captured atropine as opposed to benzatropine. The UKB participants were retained if they received any first or second generation of antipsychotics (See medication codes in Supplementary Table S3 and S4), then stratified by whether participants received any medication to treat EPSE (See EPSE medication codes in Supplementary Table S5); diagnosis of other drug-induced secondary Parkinsonism in G21.1; Drug-induced dystonia in G24.0 or Drug-induced tremor in G25.1 were selected as cases.

### GWAS Meta-analyses & Follow-up Analyses

For each set of samples, we applied logistic regression taking a within case design based on the participants’ EPSE status (participants with schizophrenia and antipsychotic exposure having EPSE vs not having EPSE) to evaluate the association between imputed SNP dosages. For UCL participants, we performed three separate GWAS for data from each wave using PLINK v2.00a2LM.^41^ The participants’ age, sex and the first three principal components of population structure were included as covariates to control for population stratification. We conducted the same sets of analyses for Aberdeen, Cardiff, and UKB samples separately where Cardiff and UKB analyses included seven additional principal components as default.

We then conducted fixed-effect meta-analysis taking each GWAS’s effective sample sizes (Neff) as weights using METAL (See calculation of Neff in Supplementary Table S6).^42^ The genome-wide significance threshold was set at P<5×10^−08^. The output results were uploaded to FUMA for interpreations.^43^

### EWAS methylation data

Methylation data was only available on a small proportion of individuals. In consideration of sample size, we switched to a between case design for the EWAS taking participants with schizophrenia and antipsychotic exposure having EPSE vs healthy controls (64 EPSE cases and 322 healthy controls from UCL; and 54 EPSE cases and 433 healthy controls from Aberdeen). In addition, comparing EPSE cases with healthy controls may provide clearer insights into accumulated methylation changes resulting from long-term antipsychotic exposure. The EZ-96 DNA Methylation kit (Zymo Research, CA, USA) was used to treat 500ng of DNA from each sample with sodium bisulfite in duplicate. DNA methylation was quantified using the Illumina Infinium HumanMethylation450 BeadChip (Illumina Inc.) run on an Illumina iScan System (Illumina) using the manufacturers’ standard protocol. Detailed data collection and imputation process has been described elsewhere.^44^ As smoking status information was not present for all samples, we estimated a proxy based on the DNA methylation profile at sites known to be associated with smoking status following a previously described approach.^45^

### EWAS analysis and Meta-analyses

As cell composition data were not available for these DNA samples, these were estimated from the DNA methylation data using both the Epigenetic Clock software^46^ and Houseman algorithm,^47,48^ including seven variables recommended in the documentation for the Epigenetic Clock in the regression analysis and smoking score. DNA methylation values for each probe were regressed against case–control status with covariates for methylation age, gender, seven cell composition scores, and smoking score. To eliminate potential schizophrenia influence from a between case design, we included the participants’ schizophrenia PRS as an additional covariate. The EWAS meta-analysis significance threshold was set at 1x10^-07^.

### EWAS Integration and Permutation test

We performed separate GWAS on the same participants used in EWAS following the same procedure described. The results were combined using METAL and clumped to represent LD independent loci in lead using the 1000 genome European samples as a reference.^49^ Any significant CpG sites from EWAS were mapped to within 250 kb of each in the associated GWAS results to identify an enrichment in the region. To quantify significance, 5000 random permutations were generated. Empirical P values for each region were calculated by counting how many of the permutations had more significant P values than the mapped P value from GWAS and dividing by the total number of permutations performed. The CpG sites’ locations were also mapped to clumped schizophrenia GWAS results within 250 kb for comparisons.^27^ Regional plots were produced using GWASLab.^50^

### PRS Calculation & Analyses

We calculated the participants’ PRSs for schizophrenia, Lewy body dementia and Parkinson’s disease using the PRS-CS method with the latest available reference GWAS.^51^ We chose the European samples from the 1000 Genomes Project Consortium as our LD reference panel given all samples included were of European Ancestry.^49^ Once weights were produced, individual PRSs were calculated using PLINK v2.00a2LM.^52^ We then used the mean and standard deviation of the healthy controls’ PRSs from each sample to standardize their cases’ PRSs. The SCZ GWAS came from Trubetskoy et al. (PGC wave 3), which were derived exclusively from European samples.^27^ The GWAS statistics for Parkinson’s disease came from European samples of Nalls et al. excluding 23andMe data.^15^ The GWAS statistics for Lewybody dementia came from Chia et al., only including European samples.^53^ We adapted the schizophrenia GWAS to exclude each sample’s participants used in the current study to avoid sample overlap. The new GWAS generation followed the same procedures as previously described.^27^

We performed multiple logistic regression analyses to assess how these various PRSs predict the presence of EPSE in each sample. Then the results were meta-analysed using a fixed effect model. The assumptions for logistic regressions were pre-checked and found to be satisfactory for each regression. The significant threshold was kept as 0.0167 (i.e. 0.05/3), for multiple testing correction.

## Results

### GWAS Sample demographics

Overall, the GWAS meta-analysis included 2471 participants with schizophrenia, of whom 1178 (48%) had EPSE. The participants had a mean age of 46.57 (SD 12.22) years old and were mostly males (70%; Table 1) as is typical of genomic studies of schizophrenia. All participants had antipsychotic exposure and most of the participants had taken at least one type of SGA (78%). The participants with and without EPSE did not differ in terms of age at assessment (46.53 vs 46.62, *p*=0.855) nor sex (males 71% vs 69%, *p*=0.213). EPSE was more prevalent in those who had taken the first generation of antipsychotics (47% vs 34%, *p*<0.001). The participants’ characteristics differed between sample sets (Supplementary Table S6). The participants who developed EPSE were at an older age at assessment than those who did not in the UCL (46.33 vs 42.72, *p*<0.001) and UKB samples (56.06 vs 53.94, *p*=0.020; Supplementary Table S6). In the Cardiff sample, participants who developed EPSE had an earlier age of schizophrenia onset (24.30 vs 27.50, *p*=0.006). The pattern of EPSE being more prevalent in those who have taken the first generation of antipsychotics were consistent across most cohorts (UCL 60% vs 40% *p*<0.001; Aberdeen 64% vs 39% *p*<0.001; UKB 69% vs 30% *p*<0.001) except for the Cardiff samples where most participants only had exposure to the second generation of antipsychotics (first 18% vs second 82%).

**Table 1.** GWAS Participants’ Demographics and Clinical Characteristics concerning EPSE Presence.

|  | N | Overall | EPSE Presence | EPSE Absence | p-values |
| --- | --- | --- | --- | --- | --- |
| **Meta-Analyses** | 2471 |  | *n = 1178 (48%)* | *n = 1293 (52%)* |  |
| **Age at assessment** | 2471 | 46.57 (12.22) | 46.53 (11.82) | 46.62 (12.59) | 0.855<sup>a</sup> |
| **Sex** | 2471 |  |  |  |  |
| Male |  | 1727 (70%) | 838 (71%) | 889 (69%) | 0.213<sup>b</sup> |
| Female |  | 744 (30%) | 340 (29%) | 404 (31%) |  |
| **Antipsychotics** | 2469 |  |  |  |  |
| First Generation |  | 989 (40%) | 554 (47%) | 435 (34%) | **<0.001<sup>b</sup>** |
| Second Generation |  | 1937 (78%) | 900 (76%) | 1037 (80%) |  |

*Notes.* EPSE, extrapyramidal side effects; SD, standard deviation
<sup>a</sup>Two Simple t-test; mean (SD)
<sup>b</sup>Pearson's Chi-squared test of independence; n (%)
In bold p passed significance threshold

### GWAS Results

We did not observe any SNP passing the genome-wide significance threshold at 5x10^-08^ (Table 2 and Supplementary Figure 1). We found no evidence for population inflation across the samples given the lambda value of 1, suggesting the test statistics are not inflated by population stratification or cryptic relatedness (Supplementary Figure 2). We observed no evidence for excessive heterogeneity across the samples. The top index SNP rs2709733 (A/G; Z=5.180; *p*=2.214×10^-07^) mapped to a long intergenic non-protein coding RNA, *LINC01162* and its effect was consistent across all cohorts (Table 2). The other affiliated protein-coding genes from the suggestive SNPs included *USP36* and *CYTH1* from rs11077391 (*p*=3.765×10^-06^) and *RBMS3* from rs6779029 (*p*=4.674×10^-06^).

### EWAS Sample demographics

The UCL participants who developed EPSE were younger than the healthy controls in terms of age at assessment (36.90 vs 44.48, *p*<0.001; See Supplementary Table S7) and methylation age (39.54 vs 44.08, *p*=0.008). The UCL EPSE cases also had higher ratios of males (81% vs 44%, *p*<0.001). The Aberdeen participants’ methylation age (54.29 vs 53.04, *p*=0.473), and males’ ratio (70% vs 74%, *p*=0.737) were balanced between the EPSE and the control group.

**Table 2.** Regions of the Genome showing the Strongest Association Signals with EPSE Presence.

| Index SNP | Chr | Position | A1/A2 | EAF | Z Score | P-values | P-values<br>Het | Match | Weight<br>(Neff) | Mapped Genes |
| --- | --- | --- | --- | --- | --- | --- | --- | --- | --- | --- |
| rs2709733 | 7 | 20878995-20955370 | A/G | 0.452 | 5.180 | $2.214 \times 10^{-07}$ | 0.378 | ++++++ | 1937 | *LINC01162* |
| rs12662039 | 6 | 99546454-99598593 | C/G | 0.057 | 4.948 | $7.492 \times 10^{-07}$ | 0.461 | +++++? | 1561 | |
| rs11077391 | 17 | 76661207-76789754 | A/G | 0.311 | 4.624 | $3.765 \times 10^{-06}$ | 0.952 | +++++? | 1561 | *USP36, CYTH1* |
| rs13175197 | 5 | 52426606-52631507 | A/C | 0.478 | 4.580 | $4.659 \times 10^{-06}$ | 0.472 | ++++++ | 1561 | |
| rs6779029 | 3 | 29026263-29110551 | T/C | 0.626 | 4.579 | $4.674 \times 10^{-06}$ | 0.950 | ?++?++ | 1530 | *RBMS3* |
| rs1817573 | 9 | 7546436-7626824 | A/G | 0.126 | 4.434 | $9.244 \times 10^{-06}$ | 0.560 | ?++++? | 1436 | |

*Notes.* Index SNP, the single-nucleotide polymorphism with the strongest association in the genomic region and each is independent at $r^2 < 0.1$ ; Chr, chromosome; Position, the start and end position (UCSC hg19) of the SNP locus where near-by SNPs were clumped to with nominal associations ( $p < 0.05$ ) and LD ( $r^2 < 0.1$ ) within 250-kb windows taking the 1000 genomes project phase 3 EUR as LD reference; A1/A2, effect and alternate allele; EAF, the effect allele frequency based on 1000 genomes EUR; Z-score, the meta-analysis output reference score for the SNP; P-values, the corresponding p-values to the candidate SNP; P-values Het, corresponding p-values to the degree of variability in effect sizes from METAL analysis; Match, the agreement across the six datasets, + means individuals who carry the A1 allele have positive EPSE association, - means negative, ? means missing, the orders are: three sets of GWAS from UCL samples (1) Affymetrix Array, (2) Illumina PsychArray, (3) GSA, Illumina Global Screening Array, (4) Aberdeen samples, (5) UKB samples, and (6) Cardiff samples; Weight, the overall Neff of the sample for the SNP; Mapped genes, the top genes mapped by positional mapping criterion with maximum distance 10kb to the locus position. No SNP passed genome-wide significant threshold.

### EWAS Meta-analysis Results and Permutation Testing

In total, we identified 9 differentially methylated positions (DMPs) associated with EPSE presence among schizophrenia participants (*p*<1×10^−07^) when controlling for methylation age, sex, derived estimates of cell composition, smoking score, and schizophrenia PRS (Table 3). Five of these identified DMPs have been implicated by past schizophrenia EWAS meta-analysis including cg12524168, p=7.61×10^-20^; cg05419385, p=3.08×10^-18^; cg22583147, p=5.66×10^-22^; cg12044923, p=1.32×10^-19^; and cg20730966, p=4.90×10^-24^.^44^ The other four DMPs cg14531564, cg20647656, cg12004641, cg22845912, and their affiliated genes *SDF4*, *ANKMY1*, *TNS1*, *SLA* were not identified in past schizophrenia or smoking EWAS.^45,54^

**Table 3.**
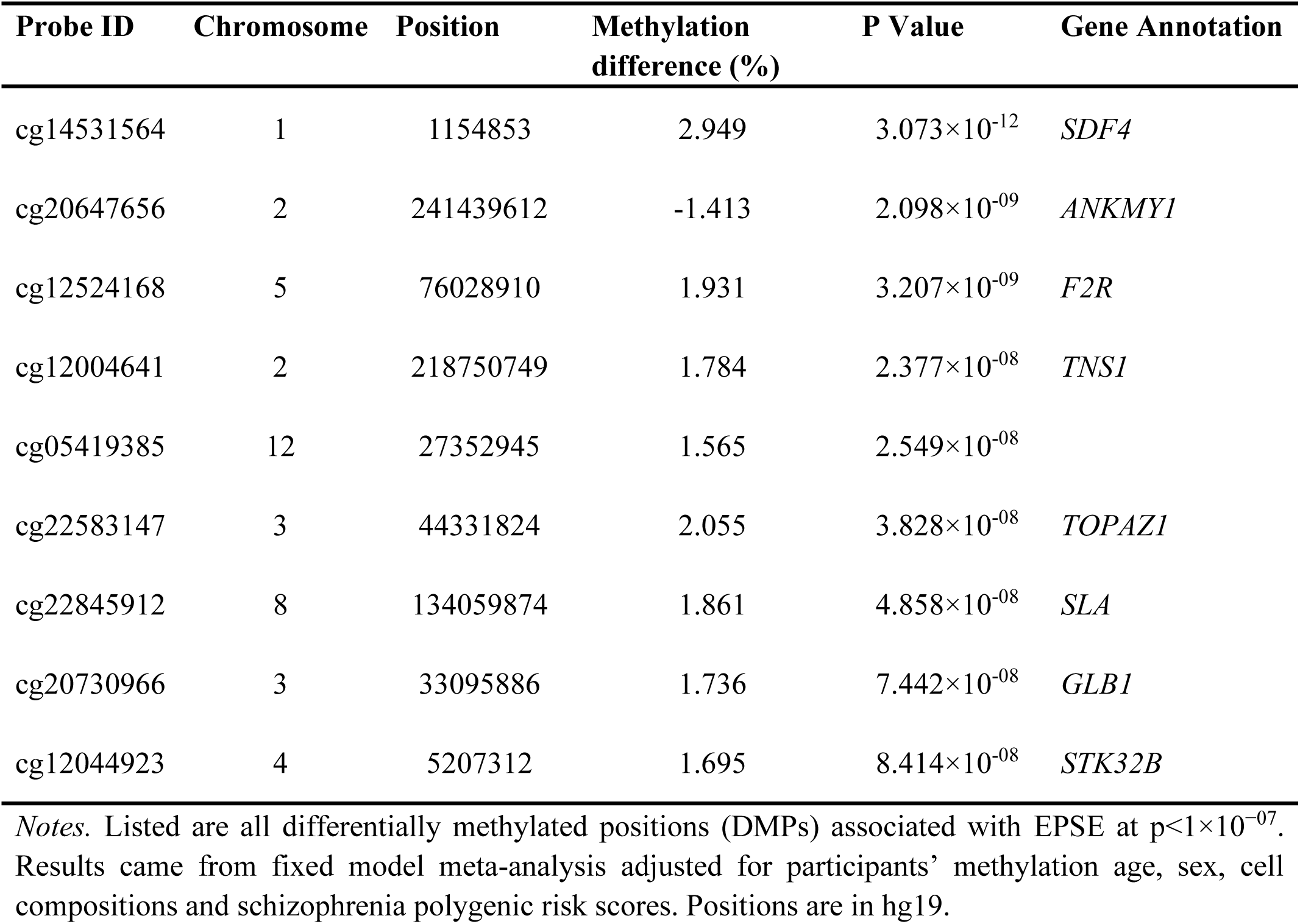
EPSE-associated differentially Methylated Positions.

We next examined whether the locations of theses DMPs could map to the corresponding GWAS of the same samples or to previously published schizophrenia GWAS. The GWAS summary statistics were first clumped so that multiple non-independent associations were collapsed into single associated loci. None of the identified DMPs were found to be associated with any genome-wide significant loci from past schizophrenia GWAS according to our regional mapping (Supplementary Figure 3-11).^27^ The SNP rs7622757 within a 250kb window with cg22583147 was closest to genome-wide significance at *p*=4.44×10^-07^ (Supplementary Figure 8).

Our mapping of the DMPs to the GWAS of associated samples found that cg12044923 was significantly associated (permutation *p*=0.010) with index SNP rs13108591 which had a GWAS p value of 7.482×10^-05^. cg20647656 was associated (permutation *p*=0.030) with index SNP rs75037293 which had a GWAS p value of 2.73×10^-04^. According to the past schizophrenia GWAS, the SNPs rs13108591 (T/C) had p value of 0.761 and rs75037293 (G/C) had p value of 0.117 indicating minor relevance to schizophrenia.^27^ The SNP rs13108591 is located on chr4:5162317 (hg19), mapping to the intron of *STK32B*. SNP rs75037293 is located on chr2:241453995 (hg19) mapping to the intron of *ANKMY1*.

### PRS Results

We found no evidence to suggest any of the selected PRS could predict the development of EPSE (Table 4). According to the fixed model meta-analysis, the participants’ genetic predisposition to Schizophrenia (*p*=0.566), Parkinson’s disease (*p*=0.492), and Lewy-body dementia (*p*=0.765) were not associated with the presence of EPSE.

**Table 4.** Results of Multiple Regression Analyses.

| Variables | Estimated Coefficient | Standard Error | Confidence Intervals (95%) | Reference Values | *p* |
| --- | --- | --- | --- | --- | --- |
| **Schizophrenia PRS** |  |  |  |  |  |
| UCL | 0.013 | 0.017 | -0.020, 0.047 | 0.780 | 0.436 |
| Aberdeen | 0.006 | 0.020 | -0.033, 0.046 | 0.321 | 0.749 |
| UKB | 0.032 | 0.017 | -0.036, 0.032 | 1.941 | 0.053 |
| Fixed-effect model | 0.019 | 0.010 | -0.001, 0.039 | 1.828 | 0.566 |
| **Parkinson's disease PRS** |  |  |  |  |  |
| UCL | 0.015 | 0.017 | -0.018, 0.049 | 0.892 | 0.373 |
| Aberdeen | 0.025 | 0.022 | -0.017, 0.068 | 1.162 | 0.246 |
| UKB | -0.010 | 0.016 | -0.042, 0.022 | -0.637 | 0.525 |
| Fixed-effect model | 0.007 | 0.010 | -0.013, 0.027 | 0.687 | 0.492 |
| **Lewy-body dementia PRS** |  |  |  |  |  |
| UCL | -0.022 | 0.016 | -0.053, 0.009 | -1.390 | 0.165 |
| Aberdeen | 0.017 | 0.023 | -0.029, 0.062 | 0.718 | 0.473 |
| UKB | 0.026 | 0.017 | -0.008, 0.060 | 1.481 | 0.139 |
| Fixed-effect model | 0.003 | 0.011 | -0.017, 0.024 | 0.299 | 0.765 |

*Notes.* PRS, polygenic risk scores
All results were adjusted for participants' age, sex, genotyping chip-type, and the first three principal components from GWAS population stratification. UKB results were adjusted for 3 additional principal components. Reference values were t values for individual models and z values for the fixed effect model.

## Discussion

In the present study, we report the largest GWAS meta-analysis of EPSE and the first EWAS meta-analysis of EPSE. The prevalence of any type of EPSE was found to be 48% among participants who have taken either FGA or SGA independent of age and sex. EPSE was found to be more prevalent among those who have taken FGA. No SNP passed the genome-wide threshold of significance. The top index SNP rs2709733 mapped to a long intergenic non-protein coding RNA, *LINC01162* with consistent effect across all cohorts. In addition, we identified multiple DMPs associated with EPSE. The gene *STK32B* implicated by cg12044923 seemed relevant to both psychiatric and movement disorder. We found no evidence that participants’ schizophrenia, Parkinson’s, and Lewy-body dementia PRSs predict EPSE development. The GWAS meta-analysis results may represent a false negative due to the limited sample size and power; but the data produced here are the result of a concerted effort to increase sample size as a starting point for future studies. Other factors may also be relevant. For example, FGA primarily target dopamine D2 receptors, particularly in the mesolimbic pathway.^6^ Strong D2 antagonism in other pathways, notably the nigrostriatal pathway, is associated with a higher risk of EPSE.^55^ SGA targets both dopamine D2 and other receptors such as *5-HT2A*. Serotonin modulation offsets some dopamine blockade effects thus reducing EPSE.^56,57^ Combining participants who have taken either FGA or SGA may conflict identification of genetic risk, given each drug type may have distinct pharmacological mechanisms and biological effects.

Alternatively, the negative GWAS and PRS findings may suggest that EPSE are more strongly driven by epigenetic modifications over time. Studies have suggested that methylation changes in dopaminergic or serotonergic pathway genes may impact motor control pathways more dynamically than SNP-based variations.^58^ This dynamic epigenetic regulation aligns with how EPSE can vary significantly among patients and change with continued antipsychotic use, whereas GWAS-derived SNPs only offer a static view of genetic risk. Therefore, integrating EWAS may provide insights into the gene-environment interactions involved in EPSE development.

Our permutation test from EWAS implicated that two genes *ANKMY1* and *STK32B* were of significant relevance to the presence of EPSE. *ANKMY1* encodes the protein Ankyrin Repeat and MYND Domain Containing 1, which has a role for protein-protein interactions and cellular signalling. This could indirectly influence pathways relevant to neurodevelopment or dopamine signalling. However, we have found little additional corroborating evidence directly linking *ANKMY1* to schizophrenia or EPSE. The other implicated gene *STK32B* encodes for a member of the human N-myristoylated proteins, which are involved in various cellular signalling and transduction pathways, although its exact biological function remains insufficiently defined.^59^ A 520-kb homozygous deletion encompassing *STK32B* has been described in Ellis-Van-Creveld syndrome, which is a rare genetic disorder that primarily affects the skeletal system and other tissues.^60^

Notably, changes in the methylation of the *STK32B* promoter region have been previously linked to both schizophrenia and anxiety disorders. It is suggested that the protein plays a role in executive functions such as working memory and selective attention.^44,61^ Moreover, *STK32B* was implicated in a GWAS of essential tremor,^62^ and patients with essential tremor showed increased expression of *STK32B* in the cerebellar cortex, highlighting a potential relevance to movement abnormalities.

The current study has several limitations. First, the study’s data collection was cross-sectional and we could not investigate how the methylation shifts were introduced by anti-psychotics and EPSE development over time. We used a mixed definition of EPSE, which may introduce variability in the classification and characterization of EPSE symptoms. Additionally, our analysis included mixed antipsychotic medications and EPSE medications, each of which may have distinct profiles of EPSE risk, contributing to potential heterogeneity in the manifestations of EPSE. This variability could impact the consistency of our findings and warrant careful consideration in future studies to clarify the effects of specific antipsychotic medications on EPSE with increased sample size to do so. In addition, although we have implemented strategies to control for collider bias related to schizophrenia, our results may still be influenced by participants’ genetic predispositions to schizophrenia.

Overall, our study provides new insights into the biological mechanisms underlying EPSE development in patients with schizophrenia. Notably, our approach integrated findings from EWAS with GWAS results, allowing us to explore EPSE- associated methylation shifts using accessible SNP data. The findings of this study indicate that further investigation of the epigenetics of EPSE and the role of *STK32B* in EPSE is likely to enhance our understanding and inform future research and treatment directions.

## Data Availability

All data produced in the present study are available upon reasonable request to the authors

## Acknowledgments

The UCL participants’ genetic and clinical data was collected with support from, the Neuroscience Research Charitable Trust, the Central London NHS (National Health Service) Blood Transfusion Service, the Camden and Islington NHS Foundation Trust, a research lectureship from the Priory Hospitals and the National Institutes for Health Research (NIHR) Mental Health Research Network (MHRN). JT was supported by NIHR HDR-UK. AM and NB were supported by the UCLH NIHR BRC. ALP was supported by a legacy donation from the Schizophrenia Association of Great Britain. SKL was funded by a PhD studentship from Mental Health Research UK (MHRUK). MJO, MCOD, and JTRW are supported by a collaborative research grant from Takeda Pharmaceuticals Ltd. for a project unrelated to work presented here. AFP, MJO, MCOD, and JTRW also reported receiving grants from Akrivia Health for a project unrelated to this collaboration. The authors have declared that there are no conflicts of interest in relation to the subject of this study.

