## Supplementary Tables 1-7 & Supplementary Figures 1-11 for "Integrating Genome-wide and Epigenome-wide Associations for Antipsychotic Induced Extrapyramidal Side Effects"

**Supplementary Table 1. Behavioural Key Words used to Identify EPSE Cases**

| Symptoms | Keywords |
|---|---|
| General terms for EPSE | Extrapyramidal; extra pyramidal; epse; EPSE; movement disorder |
| Parkinsonism (tremor) | Tremor; tremble; shake; quiver |
| Parkinsonism (rigidity) | Rigid; hypertonic; stiff; inflexible; tight; tense; muscle pain |
| Parkinsonism (Sialorrhea) | Salivation; sialorrhea; drool |
| Parkinsonism (bradykinesia and hypokinesia) | Hypokinesia; bradykinesia; freeze; froze; slow; motor block; stride; reduced arm swing; blink; hypophonia; soft voice; voice volume; slurred; micrographia; handwriting |
| Parkinsonism (mask face) | Mask; hypomimia; expressionless |
| Parkinsonism (stooped posture) | Stoop; hunch; posture |
| Parkinsonism (parkinsonian gait) | Shuffle; festinate; parkinsonian |
| Parkinsonism (parkinsonian gait) | Imbalance; fall |
| Dystonia (general) | Dystonia; jerk; twist; twitch; spasm; lock |
| Dystonia (opisthotonos) | Opisthotonos; arch back; bend back; arch spine; bend spine |
| Dystonia (torticollis) | Torticollis; cervical dystonia; head deviation; head posturing; neck pain |
| Dystonia (oculogyric crisis) | Oculogyric; fix stare; fix eye; deviate eye |
| Dystonia (trismus) | Trismus; lock jaw; lockjaw; jawlock; jaw deviation; jaw retraction; clench; grind; mouth pain; restrict mouth; limit mouth |
| Dystonia (tortipelvic crisis) | Tortipelivic; bend trunk; twist trunk |
| Dystonia (buccolingual crisis) | Buccolingual; dysphagia; difficult swallow; grimace; protrude; protrusion; risus sardonicus; dysarthria; difficult speak; difficult speech; pseudomacroglossia; swollen tounge; tounge swell |
| Dystonia (laryngeal dystonia) | Laryngeal; stridor; strangled voice; breathy voice; quiet voice; whispery voice; hoarse; shaky voice; aphonia; interrupt speech; lose voice; voice loss |
| Tardive Dyskinesia (general) | tardive dyskinesia; dyskinesia; TD |
| Tardive Dyskinesia (orofacial dyskinesia) | Involuntary; tongue twist; tongue protrusion; chew; biting; bite; suck; clench; lateral jaw movement; sideway jaw movement; smack lip; lip purse; pucker; puff cheek; frown; blink; grimace; blink |
| Tardive Dyskinesia (limb truncal dyskinesia) | Choreiform; athetoid; choreoathetoid; purposeless; rock; twist; squirm; gyrate; thrust; knee move; tap; heel drop; writhing; rotate; nod; inversion; eversion |
| Akathisia | Akathisia; akathisia; restless; pace; fidget; irritable; leg cross; leg swing; foot shift; shuffle; tramp; still; march; shift weight; rock |

**Supplementary Table 2. Pharmacological Keywords used to Identify EPSE Cases**

| **Medication** | **Keywords** |
|---|---|
| Trihexyphenidyl | Trihex; benzh; artane; agitane; parkin |
| Benzatropine | Benza;tropin; cogentin |
| Procyclidine | Procy; lidin; kemad |
| Orphenadrine | Orphan; drine |
| Biperiden | Biper; akineton |
| Hyoscine | Hyos; kwell |
| Tetrabenazine | Tetrab; ranb; nitom; xenaz |

**Supplementary Table 3. First Generation Antipsychotics used to Select Participants**

| Medication Name | Medication Type | Medication Code |
|---|---|---|
| Benperidol | anquil 250micrograms tablet; benperidol | 1140867080; 1140867078 |
| Chlorpromazine | chlorpromazine; cpz - chlorpromazine; largactil 10mg tablet; chloractil 25mg tablet | 1140879658; 1140910358; 1140863416; 1140863410 |
| Flupentixol | flupentixol; depixol 3mg tablet; fluanxol 500micrograms tablet; flupenthixol; flupentixol | 1140909800; 1140867152; 1140867952; 1140867150; 1140909800 |
| Fluphenazine | decazate 25mg/1ml oily injection; fluphenazine; fluphenazine decanoate; modecate 12.5mg/0.5ml oily injection; moditen 1mg tablet; moditen enanthate 25mg/ml injection | 1140867474; 1140882098; 1140867398; 1140867456; 1140867156; 1140856004 |
| Haloperidol | haldol 5mg tablet; haloperidol; serenace 500micrograms capsule | 1140867184; 1140867168; 1140867092 |
| Levomepromazine | levomepromazine; nozinan 25mg tablet | 1140909802; 1140867122 |
| Loxapine | loxapine; loxapac 10mg capsule | 1140867406; 1140867414 |
| Pericyazine | neulactil 2.5mg tablet; pericyazine | 1140867136; 1140867134 |
| Perphenazine | fentazin 2mg tablet; perphenazine | 1140867210; 1140867208 |
| Pimozide | orap 2mg tablet; pimozide | 1140867272; 1140867218 |
| Pipotiazine | piportil depot 50mg/1ml oily injection; pipotiazine | 1140867572; 1140909804 |
| Prochlorperazine | prochlorperazine; stemetil 5mg tablet | 1140868170; 1140868172 |
| Promazine | promazine | 1140879746 |
| Sulpiride | dolmatil 200mg tablet; sulparex 200mg tablet; sulpiride; sulpitil 200mg tablet; sulpor 200mg/5ml oral solution | 1140867306; 1140917366; 1140867304; 1140882376; 1141185130 |
| Thioridazine | thioridazine; melleril 10mg tablet | 1140879750; 1140867312 |
| Trifluoperazine | stelazine 1mg tablet; tranylcypromine+trifluoperazine 10mg/1mg tablet; trifluoperazine | 1140867244; 1140867944; 1140868120 |
| Zuclopenthixol | clopixol 2mg tablet; zuclopenthixol | 1140867342; 1140882100 |

**Supplementary Table 4. Second Generation Antipsychotics used to Select Participants**

| Medication Name | Medication Type | Medication Code |
|---|---|---|
| Amisulpride | amisulpride; solian 100mg/ml s/f oral solution | 1141153490; 1141184742 |
| Aripiprazole | abilify 5mg tablet; aripiprazole | 1141202024; 1141195974 |
| Clozapine | clozapine; clozaril 25mg tablet; denzapine 25mg tablet | 1140867420; 1140882320; 1141200458 |
| Olanzapine | olanzapine; zyprexa 2.5mg tablet | 1140928916; 1141167976 |
| Oxypertine | oxypertine; integrin 10mg capsule | 1140879754; 140855978 |
| Quetiapine | quetiapine; seroquel 25mg tablet | 1141152848; 1141152860 |
| Remoxipride | remoxipride; roxiam 150mg m/r capsule | 1140879704; 1140867432 |
| Risperidone | dozic 1mg/ml oral liquid; risperdal 0.5mg tablet; risperidone | 1140867180; 1141177762; 1140867444 |
| Sertindole | serdolect 4mg tablet; sertindole | 1140927970; 1140927956 |

**Supplementary Table 5. EPSE Medications used to Select Cases**

| Medication Name | Medication Type | Medication Code |
|---|---|---|
| Akineton | akineton | 1140872522 |
| Artane | artane | 1140872378 |
| Benzatropine | benzatropine | 1140909818 |
| Benzhexol | benzhexol | 1140883510 |
| Biperiden | biperiden | 1140872520 |
| Cogentin | cogentin | 1140872460 |
| Kemadrin | kemadrin | 1140872542 |
| Orphenadrine | orphenadrine | 1140883560 |
| Procyclidine | procyclidine | 1140883476 |
| Tetrabenazine | tetrabenazine | 1140872556 |
| Tetrabenazine Product | tetrabenazine_product | 1141157336 |
| Trihexyphenidyl | trihexyphenidyl | 1140909816 |
| Xenazine | xenazine | 1141171726 |

**Supplementary Table 6. GWAS Participants' Demographics and Clinical Characteristics concerning EPSE Presence**

| | N | Overall | EPSE Presence | EPSE Absence | Neff | p-values |
|---|---|---|---|---|---|---|
| **UCL** | *1017* | | *n = 587 (58%)* | *n = 430 (42%)* | *983* | |
| **Age at assessment** | 1017 | 44.80 (12.24) | 46.33 (12.10) | 42.72 (12.14) | | **<0.001**[a] |
| **Age of onset** | 761 | 23.34 (8.18) | 23.28 (8.07) | 23.44 (8.34) | | 0.795[a] |
| **Sex** | 1017 | | | | | 0.145[b] |
| Male | | 735 (72%) | 435 (74%) | 300 (70%) | | |
| Female | | 282 (28%) | 152 (26%) | 130 (30%) | | |
| **Antipsychotics** | 973 | | | | | **<0.001**[b] |
| First generation | | 521 (54%) | 351 (60%) | 170 (40%) | | |
| Second generation | | 871 (90%) | 482 (82%) | 389 (90%) | | |
| **Aberdeen** | *414* | | *n = 90 (22%)* | *n = 324 (78%)* | *282* | |
| **Age at assessment** | 414 | 44.40 (13.20) | 45.70 (13.12) | 44.03 (13.21) | | 0.292[a] |
| **Age of onset** | 401 | 24.08 (8.05) | 23.27 (7.76) | 24.29 (8.13) | | 0.293[a] |
| **Sex** | 414 | | | | | 0.426[b] |
| Male | | 311 (75%) | 71 (79%) | 240 (74%) | | |
| Female | | 103 (25%) | 19 (21%) | 84 (26%) | | |
| **Antipsychotics** | 414 | | | | | **<0.001**[b] |
| First generation | | 183 (44%) | 58 (64%) | 125 (39%) | | |
| Second generation | | 279 (67%) | 48 (53%) | 231 (71%) | | |
| **UKB** | *507* | | *n = 90 (18%)* | *n = 417 (82%)* | *296* | |
| **Age at assessment** | 507 | 54.32 (8.08) | 56.06 (7.61) | 53.94 (8.14) | | **0.020**[a] |
| **Sex** | 507 | | | | | 0.322[b] |
| Male | | 335 (66%) | 64 (71%) | 271 (65%) | | |
| Female | | 172 (34%) | 26 (29%) | 146 (35%) | | |
| **Antipsychotics** | 507 | | | | | **<0.001**[b] |
| First generation | | 187 (37%) | 62 (69%) | 125 (30%) | | |
| Second generation | | 349 (69%) | 38 (42%) | 311 (75%) | | |
| **Cardiff** | *533* | | *n = 411 (77%)* | *n = 122 (12%)* | *376* | |
| **Age at assessment** | 533 | 44.30 (11.70) | 44.90 (10.90) | 42.20 (14.10) | | 0.057[a] |
| **Age of onset** | 508 | 25.00 (8.80) | 24.30 (7.60) | 27.50 (11.60) | | **0.006**[a] |
| **Sex** | 533 | | | | | 0.900[b] |
| Male | | 346 (65%) | 268 (65%) | 78 (64%) | | |
| Female | | 187 (35%) | 143 (35%) | 44 (36%) | | |
| **Antipsychotics** | 531 | | | | | 0.100[b] |
| First generation | | 98 (18%) | 83 (20%) | 15 (12%) | | |
| Second generation | | 438 (82%) | 332 (81%) | 106 (87%) | | |

*Notes.* EPSE, extrapyramidal side effects; SD, standard deviation

[a] Two Simple t-test; mean (SD)

[b] Pearson's Chi-squared test of independence; n (%)

In bold p passed significance threshold

Neff, effective sample sizes, calculated as 4/(1/n_cases + n_controls); UCL Neff is a sum of Neff from 3 separate genotyping waves (Affymetrix 125; PsychArray 579; Global Screening Array 279)

**Supplementary Figure 1. The Manhattan Plot of EPSE GWAS Meta-analysis**

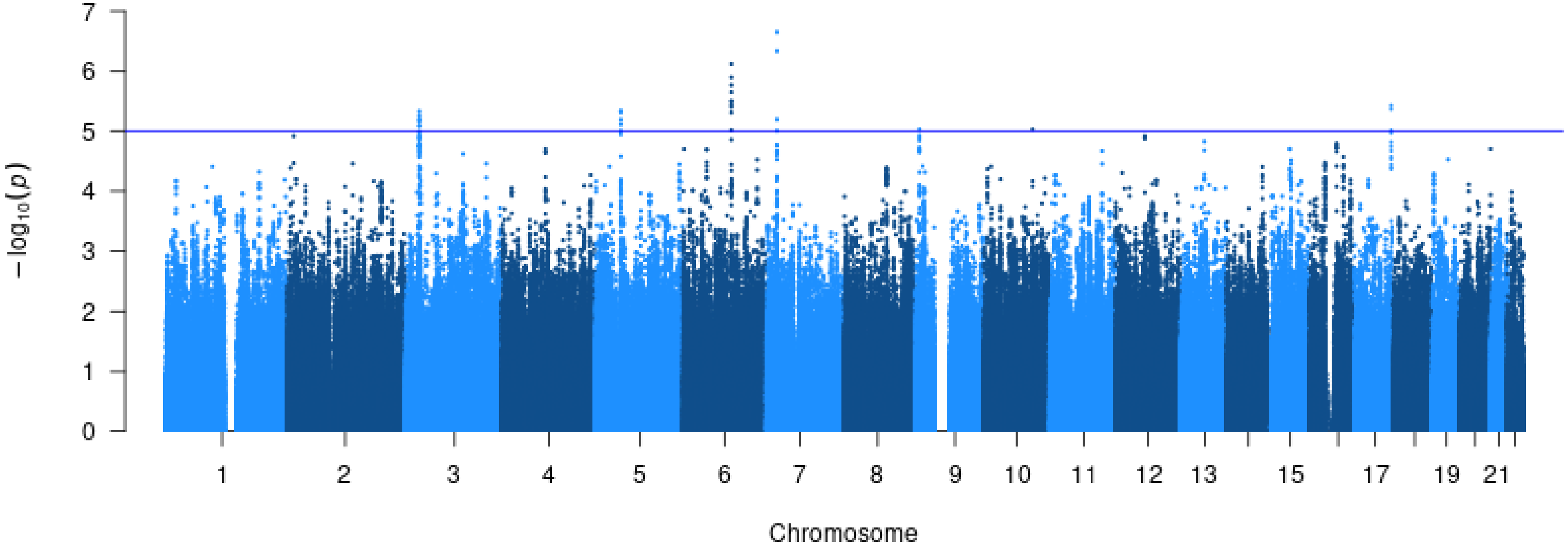

**Supplementary Figure 2. The QQ Plot of EPSE GWAS Meta-analysis**

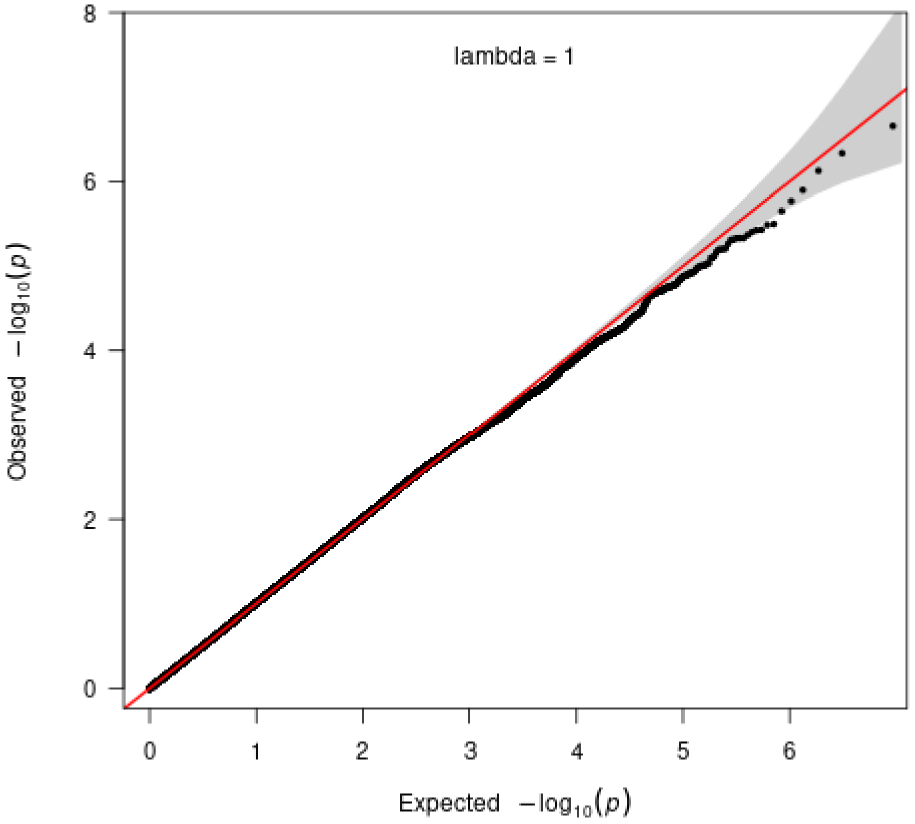


*Notes.* X-axis (Expected -log10(p-values)): Represents the theoretical quantiles under the null hypothesis, where p-values are uniformly distributed on a logarithmic scale; Y-axis (Observed -log10(p-values)): Shows the observed -log10(p-values) from the GWAS.

**Supplementary Table 7. EWAS Participants' Demographics and Clinical Characteristics concerning EPSE Presence**

| | N | Overall | EPSE Presence | Controls | p-values |
|---|---|---|---|---|---|
| **UCL** | 379 | | n = 64 (17%) | n = 315 (83%) | |
| **Age at assessment** | 362 | 38.24 (14.81) | 36.90 (14.74) | 44.48 (13.58) | **<0.001**[a] |
| **mAge (Horvath)** | 379 | 40.31 (12.46) | 39.54 (12.41) | 44.08 (12.13) | **0.008**[a] |
| **Sex** | | | | | **<0.001**[b] |
| Male | | 192 (51%) | 52 (81%) | 140 (44%) | |
| Female | | 187 (49%) | 12 (19%) | 175 (56%) | |
| **Antipsychotics** | 64 | | | | |
| First generation | | | 57 (89%) | / | |
| Second generation | | | 50 (78%) | / | |
| **Aberdeen** | 480 | | n = 47 (10%) | n = 433 (90%) | |
| **mAge (Horvath)** | 480 | 53.16 (9.88) | 54.29 (11.48) | 53.04 (9.70) | 0.473[a] |
| **Sex** | 480 | | | | |
| Male | | 352 (73%) | 33 (70%) | 319 (74%) | 0.737[b] |
| Female | | 128 (27%) | 14 (30%) | 114 (36%) | |
| **Antipsychotics** | 47 | | | | |
| First generation | | | 30 (64%) | / | |
| Second generation | | | 23 (49%) | / | |

*Notes.* EPSE, extrapyramidal side effects; SD, standard deviation

[a] Two Simple t-test; mean (SD)

[b] Pearson's Chi-squared test of independence; n (%)

In bold p passed significance threshold

Supplementary Figures 3–11 describe mapping of significant CpG sites to Schizophrenia GWAS within a 250kb window. Black line in the central mark the SNP closest to the CpG. Red dotted line marked the SNP in lead within the region and other nearby SNPs in linkage disequilibrium.

**Supplementary Figure 3. The Mapping of CpG cg14531564 to the Schizophrenia GWAS Region**

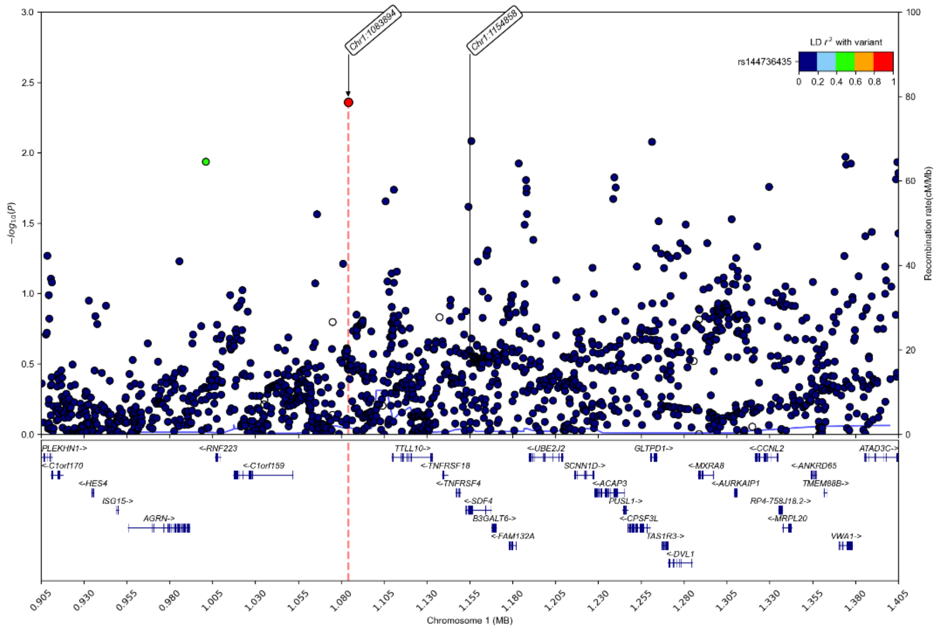

### Supplementary Figure 4. The Mapping of CpG cg20647656 to the Schizophrenia GWAS Region

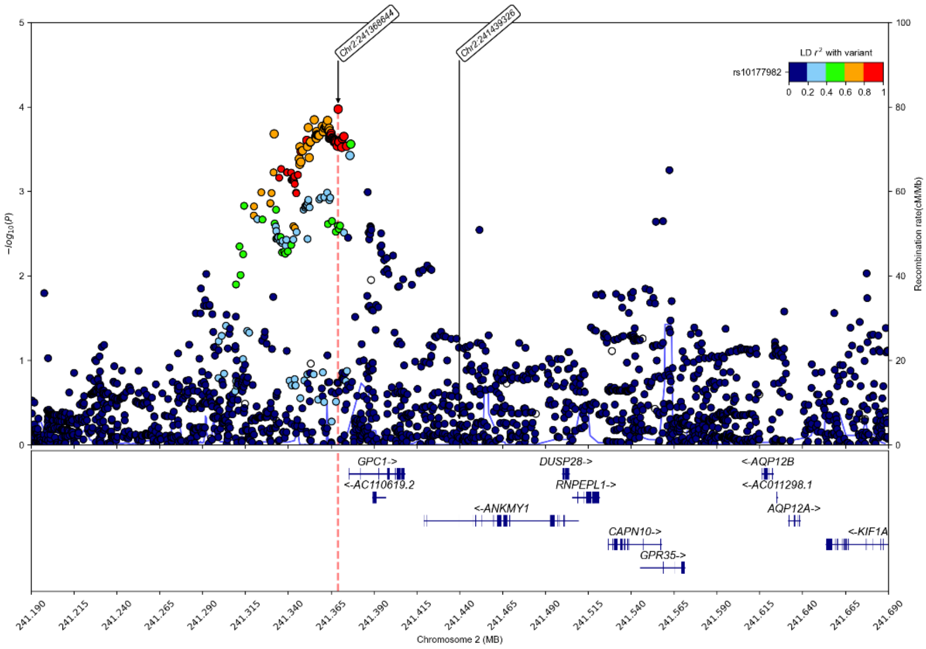


### Supplementary Figure 5. The Mapping of CpG cg12524168 to the Schizophrenia GWAS Region

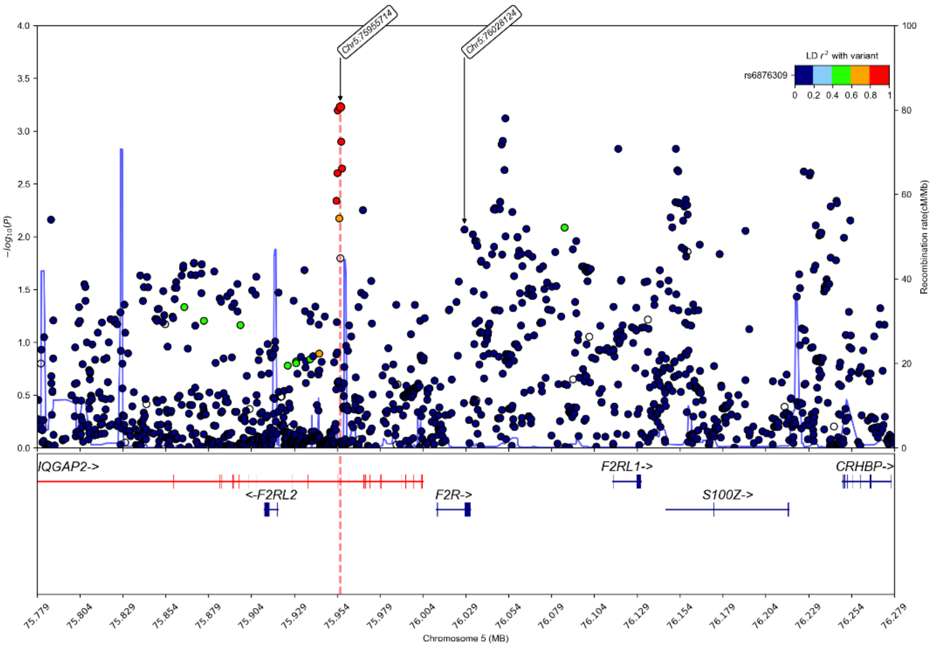

**Supplementary Figure 6. The Mapping of CpG cg12004641 to the Schizophrenia GWAS Region**

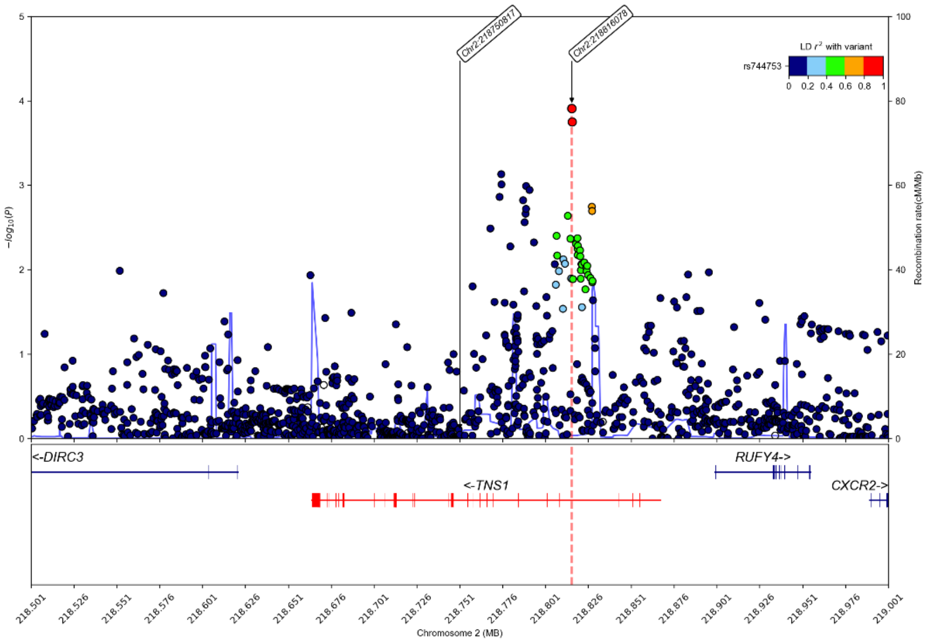


**Supplementary Figure 7. The Mapping of CpG cg05419385 to the Schizophrenia GWAS Region**

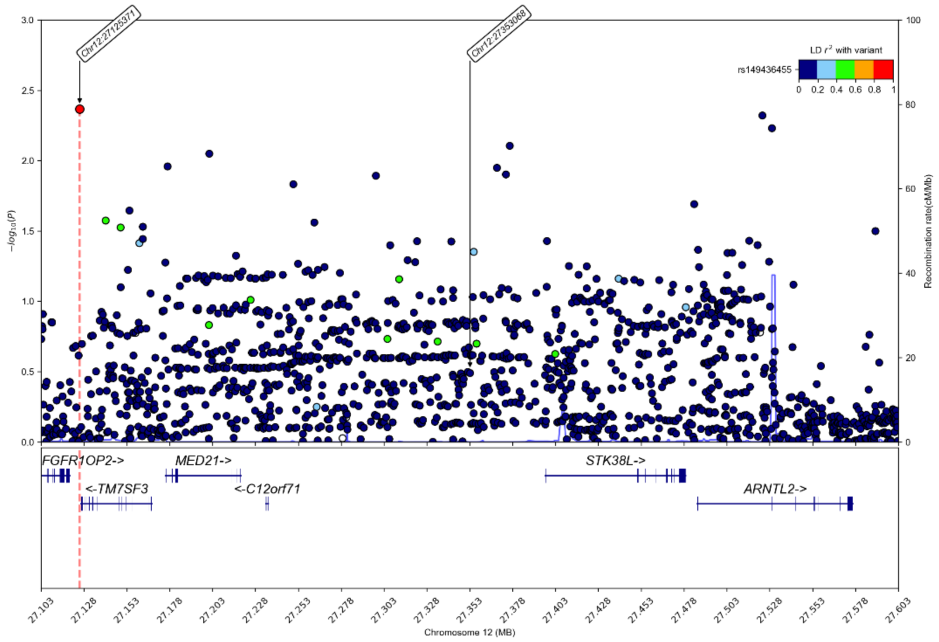

**Supplementary Figure 8. The Mapping of CpG cg22583147 to the Schizophrenia GWAS Region**

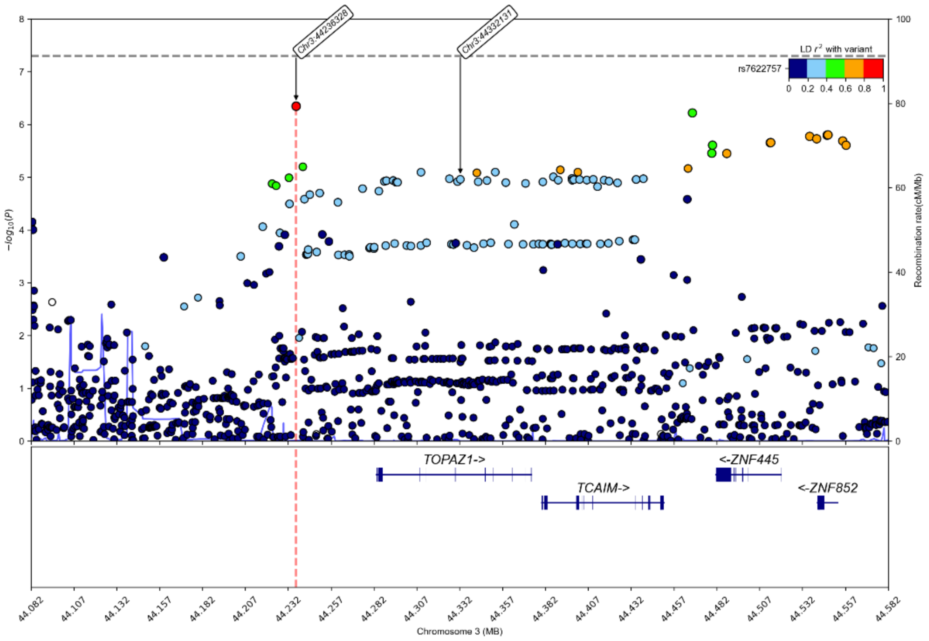


**Supplementary Figure 9. The Mapping of CpG cg22845912 to the Schizophrenia GWAS Region**

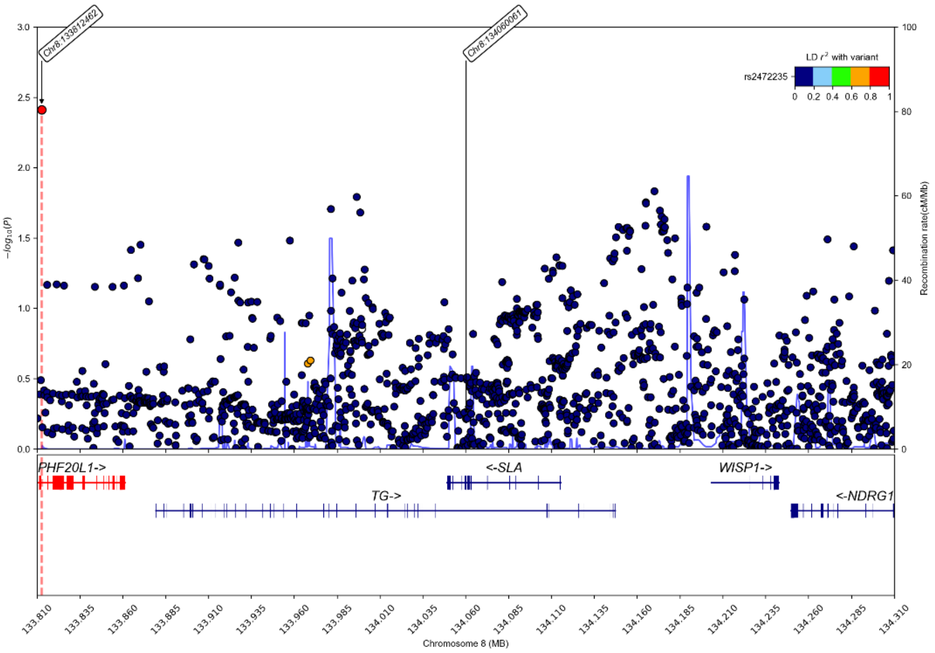

### Supplementary Figure 10. The Mapping of CpG cg20730966 to the Schizophrenia GWAS Region

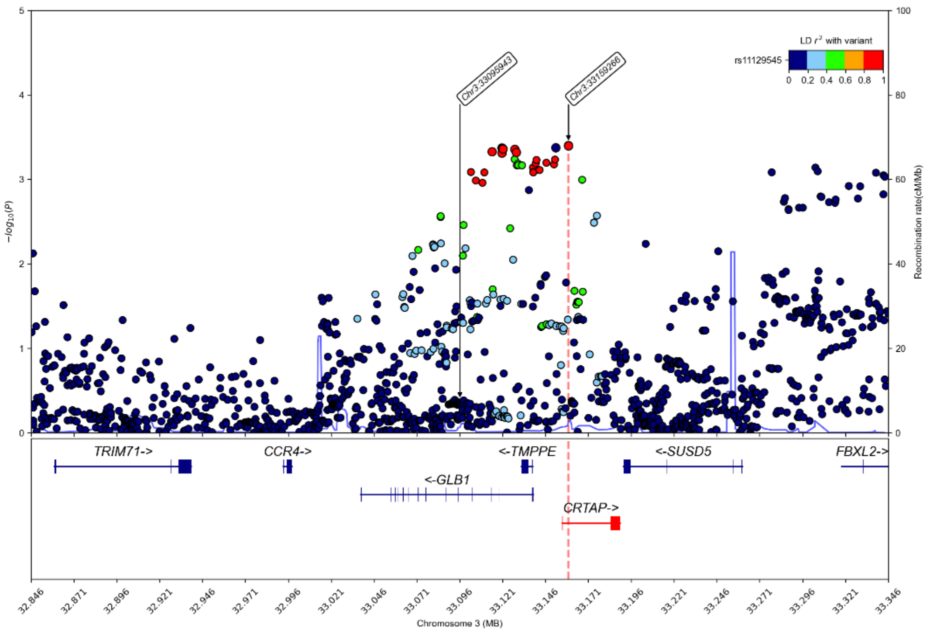


### Supplementary Figure 11. The Mapping of CpG cg12044923to the Schizophrenia GWAS Region

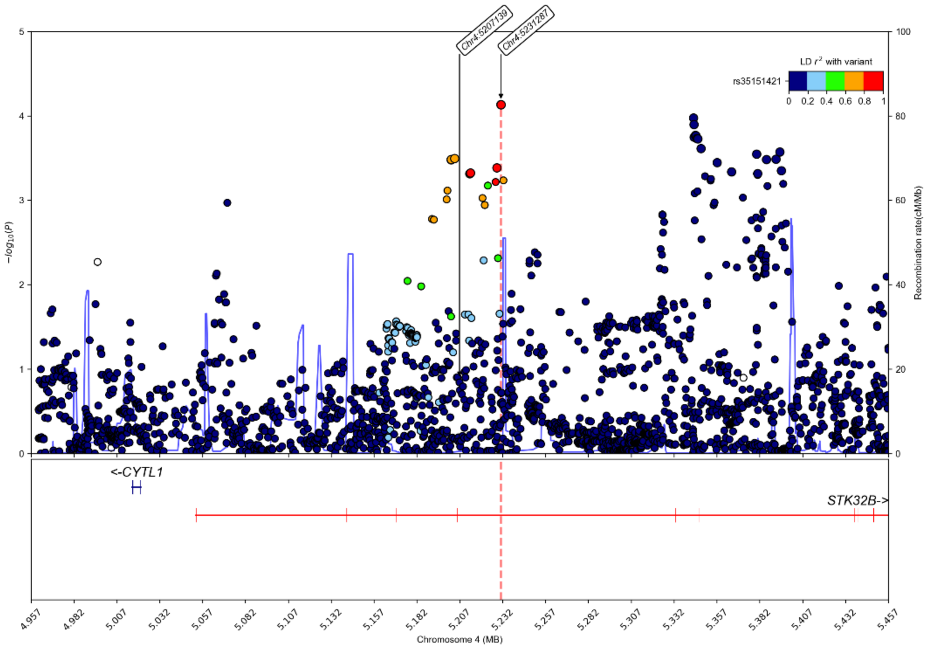
